# Clinical indicants and electrophysiological patterns of sciatic nerve in patients after total hip replacement

**DOI:** 10.1101/2021.10.11.21264505

**Authors:** V.V. Ostrovskij, V.S. Tolkachev, S.P. Bazhanov, G.A. Korshunova, A.A. Chekhonatsky

## Abstract

**Background:** The incidence of sciatic nerve (SN) damage after a total hip replacement (THR) is 10 percent. The underdiagnosis of paucisymptomatic sciatic neuropathy may lead to the unsatisfactory outcome of the treatment in these patients featured by frequent chronic pain syndrome. This research **was aimed at** the evaluation of the dynamics of clinical and electrophysiological patterns in SN after THR.

**Material and methods:** The research involved 16 individuals 45 to 68 years old with primary idiopathic coxarthrosis who underwent THR in the Scientific Research Institute of Traumatology, Orthopedics and Neurosurgery, Federal State Budgetary Educational Institution of Higher Education ‘V.I. Razumovsky Saratov State Medical University’, the Russian Federation Ministry of Healthcare. We compared the findings of clinical and neurologic examinations (VAS, muscle strength, and sensitivity evaluation) as well as ENMG before surgeries and 14 days after them.

**Results:** In the evaluation of the clinical score in 9 patients, we observed some negative changes featured by paresthesia around the area of the SN innervation. The analysis of changes in ENMG findings revealed the decrease in M-response amplitudes of both peroneal and tibial nerves by more than 10 percent of the age-appropriate normal value; this was more prominent in patients who had undergone the lengthening of extremities in more than 3 cm.

**Conclusion:** THR is associated with a higher risk of traction and entrapment changes in SN that lead to the progress of their neuropathies in the post-operative period.

**Reviewers:** Assoc. Prof. Ulyanov V.Yu., MD, DSc;

Assoc. Prof. Gulyaev D.A., MD, DSc

## BACKGROUND

Some researchers claim the incidence of sciatic nerve (SN) damage is as high as 5 percent after a primary total hip replacement (THR) [1, 2]. The progress of SN neuropathy in THR adversely affects both the functions of the lower extremities and the mental health of the patients. The treatment of these injuries takes longer and often fails. In more than 50 percent of cases, the expressed movement deficiency that led to permanent incapacitation remained [3, 4]. However, the transient and mild disorders of SN conduction featured by morphology signs corresponding to neuropraxis (Seddon, H.J., 1943) are not uncommon. If this condition is left undiagnosed, the post-surgery management of the patient gets hindered leading to frequent chronicity of pain syndromes refractory to medical treatment [5].

The SN is formed with L4-S3 segments of the lumbosacral plexus and runs under the tendon of the piriform muscle along the posterior hip surface. However, SN anatomy varies; e.g., in 10-15 percent of cases some fibers of the nerve running through the piriform muscle and others – under its tendon [6] thus predetermining the nerve damage in THR mostly due to direct exposure particularly if the joint posterior approaches were used (Moore, Southern) [7-9].

As the published studies claim, the damage to the peroneal portion of SN is more common. This is probably due to its lateral position, a smaller number of fibrous connective fibers within the nerve structure, and the formation of the tunnel on caput fibulae level [9], all these are possible predictors of further entrapment and traction damage. It is worth noting that SN features limited tolerance to stretching, therefore post-surgery SN neuropathies after the extremity lengthening over 3 cm are frequent [10-12]. The combination of these two damage mechanisms may be also explained by coping with functional walking asymmetry in the extremities before surgery that results in compensational and adjustment responses involving the lumbar spine and leading to the progress of degenerative changes in it, or entrapment damage to nerve roots. This then causes damage to neurotrophic maintenance of effectors with dystrophic processes progressing in them [10].

Therefore, the diagnostic criteria of paucisyptomatic post-surgery neuropathy and its early detection remain underinvestigated. These challenging issues require further researching. This research had the evaluation of the dynamics of clinical and electrophysiological patterns in SN after THR as its **objective**.

## MATERIAL AND METHODS

This prospective single-center study involved 16 patients 45 to 68 years old who underwent treatment in the Scientific Research Institute of Traumatology, Orthopedics and Neurosurgery, Federal State Budgetary Educational Institution of Higher Education ‘V.I. Razumovsky Saratov State Medical University’, the Russian Federation Ministry of Healthcare from Jan. 01 to Apr. 30, 2021. The research complied with the Declaration of Helsinki principles and was approved by the local ethics committee; the authors confirm that they obtained the patients’ informed consent to participate.

All patients suffered from primary idiopathic coxarthrosis and had THR performed on them. The eligibility criteria were stage 3 primary idiopathic coxarthrosis, body-weight index from 18.50 to 24.99. The exclusionary criteria included polyneuropathy of any etiology, revision surgeries on their hips, profuse intraoperative hemorrhage (over 15 percent of BV).

All surgeries were performed only through an anterolateral approach to the hip joint, and cementless implants were used.

All patients underwent dynamic neurological examination using a visual analogue scale of pain assessment (VAS, 1974), muscle strength, and lower extremity sensitivity scales. Neuromonitoring was performed in Dantec Keypoint (Alpine Biomed ApS, Denmark) before the surgeries and in 14 days after them, and involved electroneuromyography (ENMG) of tibial and peroneal nerves and defining M-responses features. Intramuscular electromyography (iEMG) was employed to define the denervation activity of effectors for tibial, peroneal, and sciatic nerves. We estimated the fibrillation potentials and positive sharp waves.

The syndrome of SN damage was detected when the neurological examination revealed the decrease in muscle strength and sensitivity by more than 1 score of pre-surgery findings. It was also considered proven by ENMG findings if the evoked responses decreased or denervation activity in lower extremities was detected.

All patients were verticalized on day 2 of their surgeries and had to use crutches as additional support. They received combination medical (analgesics, antibiotics, anticoagulants, fluid infusions) and recovery treatment during their hospital stay.

The findings were processed with Microsoft Office Excel 2019 and IBM SPSS Statistics v23 software. Since the distribution of data did not correspond to the normal distribution law, the findings were estimated by the descriptive and nonparametric statistical methods, and Me (Q1; Q3) was also found.

## RESULTS

Before their surgeries, all patients (n=16) had severe pain syndromes in their hip areas corresponding to 8.0 (6.0; 9.0) VAS scores. The scores of the muscle strength and sensitivity in the affected extremity did not differ from those in their healthy sides and corresponded to 5 scores. No vegetative and trophic skin disorders were found in the affected lower extremity of all patients.

The pre-surgery ENMG revealed a slight decline (less than 10 percent of the age-appropriate normal value) in M-responses from peroneal and tibial nerves in 3 patients with the shortening of their lower extremities by more than 3 cm. In other cases (n=13) the amplitudes of M-responses corresponded to the age-appropriate norm. No patient featured any denervation activity through the lower leg and hip iEMG. Table 1 contains indicants (Me (Q1; Q3)) of M-responses from peroneal and tibial nerves in patients during their pre-operative period.

**Table 1.**
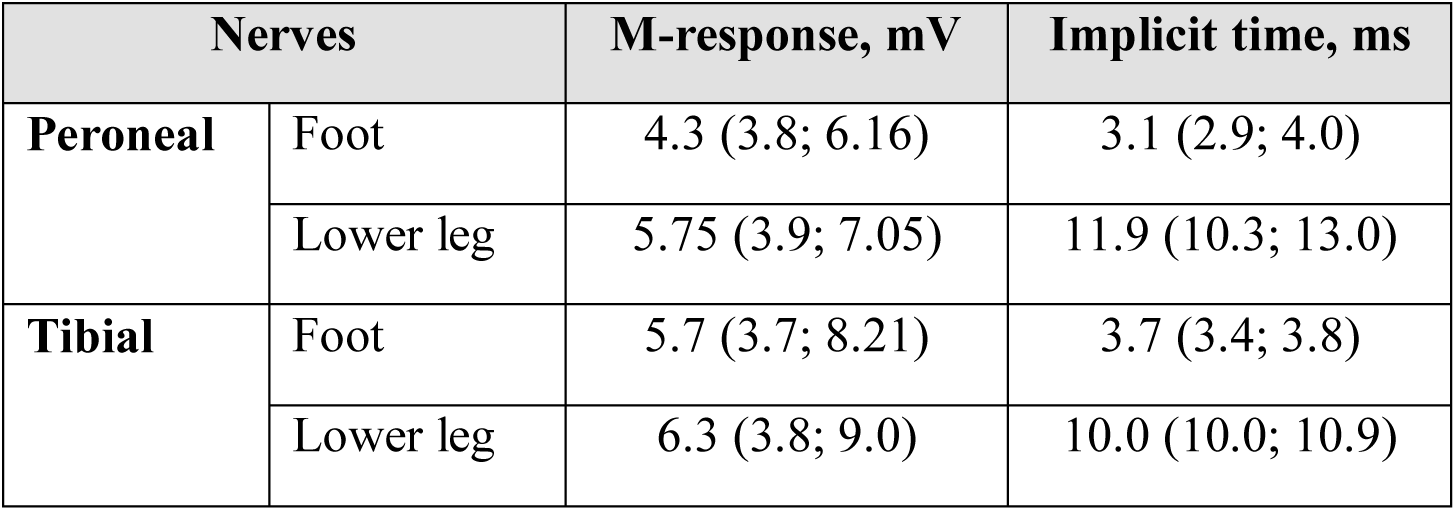

| Nerves |  | M-response, mV | Implicit time, ms |
| --- | --- | --- | --- |
| **Peroneal** | Foot | 4.3 (3.8; 6.16) | 3.1 (2.9; 4.0) |
|  | Lower leg | 5.75 (3.9; 7.05) | 11.9 (10.3; 13.0) |
| **Tibial** | Foot | 5.7 (3.7; 8.21) | 3.7 (3.4; 3.8) |
|  | Lower leg | 6.3 (3.8; 9.0) | 10.0 (10.0; 10.9) |

As Table 1 suggests, the median pre-surgery findings of peroneal and tibial M-responses in patients corresponded to the norm.

The repeated examination on day 14 of their surgeries revealed the decrease in pain syndrome to 4.0 (3.0; 5.0) VAS scores in all patients (n=16). No patients featured augmentation of movement disorders in their lower extremities. However, the sensitivity assessment revealed its decrease in the autonomic area of innervation of tibial SN portion in 4 patients and the area of innervation of both peroneal and tibial SN portions to 4 scores in 5 patients.

The follow-up ENMG revealed the decrease in M-responses of both peroneal and tibial nerves which were proven to be statistically significant (p=0.032). The worst decrease in M-response amplitude was found in patients who underwent the lengthening of their lower extremities. iENG revealed no dynamics as compared to pre-surgery findings.

Table 2 contains indicants of M-responses from peroneal and tibial nerves in patients on post-surgery day 14.

**Table 2.**

| Nerves |  | M-response, mV | Implicit time, ms |
| --- | --- | --- | --- |
| **Peroneal** | Foot | 3.9 (3.3; 5.2) | 3.2 (2.5; 6.4) |
|  | Lower leg | 5.2 (3.35; 6.42) | 10.7 (7.3; 11.52) |
| **Tibial** | Foot | 5.1 (3.0; 7.7) | 3.9 (3.5; 5.0) |
|  | Lower leg | 6.1 (3.0; 8.4) | 10.5 (7.4; 13.3) |

As Table 2 shows, the indicants of post-surgery M-response amplitudes decreased suggesting the slight syndrome of SN conduction defect.

## DISCUSSION

Our research describes the dynamics of SN ENMG in patients after primary THR. It is worth noting that none of the patients had any clinical signs of SN neuropathy in their post-surgery period but some mild sensitivity disorders. However, ENMG revealed a decrease in M-response amplitude indicants and an increase in its implicit time for the affected extremity. The changes were more prominent in those patients who had their lower extremities shortened and underwent their lengthening; this could be explained by the traction nature of the damage.

The progress of post-surgery SN neuropathy may also be associated with the entrapment nature of the damage when SN gets strained at the posterior edge of the acetabulum, and the trunk of the common peroneal nerve at fibula condyles and neck.

## CONCLUSION

Therefore, post-surgery SN neuropathy after THR is a challenging issue of multidisciplinary nature, its solving requires the participation of not just trauma orthopedists but neurologists, neurosurgeons, functional diagnostics experts, and rehabilitation therapists.

## Data Availability

The authors confirm that the data supporting the findings of this study are available within the article and/or its supplementary materials.

